# Automatic Lymph Nodes Segmentation and Histological Status Classification on Computed Tomography Scans Using Convolutional Neural Network

**DOI:** 10.1101/2024.05.07.24304092

**Authors:** Alexey Shevtsov, Iaroslav Tominin, Vladislav Tominin, Vsevolod Malevanniy, Yury Esakov, Zurab Tukvadze, Andrey Nefedov, Piotr Yablonskii, Pavel Gavrilov, Vadim Kozlov, Mariya Blokhina, Elena Nalivkina, Victor Gombolevskiy, Yuriy Vasilev, Mariya Dugova, Valeria Chernina, Olga Omelyanskaya, Roman Reshetnikov, Ivan Blokhin, Mikhail Belyaev

**Affiliations:** IRA Labs, Moscow, Russia, Skolkovo Innovation Centre territory, 30, Bolshoi boulevard, bld. 1; Moscow City Clinical Oncological Hospital № 1, Moscow, Russia, Kolomenskij proezd, d.4; Saint-Petersburg State Research Institute of Phthisiopulmonology of the Ministry of Healthcare of the Russian Federation, Saint-Petersburg, Russia, Ligovskij pr., d.2-4; Novosibirsk Regional Clinical Oncology Dispensary, Novosibirsk, Russia, ul.Plaxotnogo, dom 2; AstraZeneca Pharmaceuticals LLC, Moscow, Russia, 1-j Krasnogvardejskij pr-d, d. 21 str. 1; State Budget-Funded Health Care Institution of the City of Moscow “Research and Practical Clinical Center for Diagnostics and Telemedicine Technologies of the Moscow Health Care Department”, 127051, Moscow, Russia, Petrovka str., 24, p. 1

**Keywords:** Lung Cancer, Lymph Nodes, Medical Imaging, Deep Learning

## Abstract

Lung cancer is the second most common type of cancer worldwide, making up about 20% of all cancer deaths with less than 10% 5-year survival rate for the very late stage. The recent guidelines for the most common non-small-cell lung cancer (NSCLC) type recommend performing staging based on the 8th edition of TNM classification, where the mediastinal lymph node involvement plays a key role. However, most of the non-invasive methods have a very limited level of sensitivity and are relatively accurate, but invasive methods can be contradicted for some patients. Current advances in Deep Learning show great potential in solving such problems. Still, most of these works focus on the algorithmic side of the problem, not the clinical relevance. Moreover, none of them addressed individual lymph node malignancy classification problem, restricting the indirect analysis of the whole study, and limiting the interpretability of the result without giving an option for cliniciansto validate the result. This work mitigates these gaps, proposing a multi-step algorithm for each visible mediastinal lymph node segmentation and assessing the probability of its involvement in themetastatic process, using the results of histological verification on training. The developed pipelineshows 0.74 ± 0.01 average Recall with 0.53 ± 0.26 object Dice Score for the clinically relevant lymph nodes segmentation task and 0.73 ROC AUC for patient’s N-stage prediction, outperformingtraditional size-based criteria.

## 1 Introduction

Lung cancer is the second most common type of cancer worldwide. For the year 2018, global incidence and mortality rates were 2.1 million and 1.8 million, respectively, making about 20% of all cancer deaths Thandra et al. [2021]. The five-year survival rate prognosis starts from 68-92% for the early stage and dramatically drops below the level of 10% for the very late stage, which is 42% of all cases Goldstraw et al. [2016]. So, it is crucial to indicate and treat lung cancer early to increase the patient’s survival chances and lower the treatment process costs.

The worldwide practice for indicating lung cancer in the early stages is screening programs. These are examinations for finding a disease among the group of risk patients without any symptoms. The 50+ y.o., current (or quit for the last 15 years) smokers, having at least a 20-pack-year smoking history, are recommended to participate in such programs Tanoue et al. [2015]. The recommended Ettinger et al. [2019], Planchard et al. [2018] method to be used for lung screening programs is a low-dose computed tomography (LDCT), the efficacy of which was proven by several randomized prospective studies Heleno et al. [2018], Pegna et al. [2013], Infante et al. [2015], de Koning et al. [2020],Pastorino et al. [2019], Baldwin et al. [2011]. Despite being non-invasive and minimally harmful, it doesn’t provide sufficient insights needed for incident-finding verification.

For the initial finding verification, the guidelines Ettinger et al. [2019], Planchard et al. [2018] recommend using non-invasive positron emission tomography combined with computed tomography (PET-CT) scanning procedures or minimally-invasive biopsy. Once the initial finding is verified as cancer, the clinicians have to understand its spread (this process is called “staging”) and determine the best treatment tactics, depending on the lung cancer type.

The recent guidelines for the most common non-small-cell lung cancer (NSCLC) type recommend performing staging based on the 8th edition of TNM classification Detterbeck et al. [2017]. The T, N, and M letters stand for the evaluated entities. Primary tumor, thoracic lymph nodes, and distant metastases in other organs, respectively. The combination yields the patient’s stage that can be clinical (usually done before the surgery using non-invasive methods), pathological (using histological analysis), and re-staging after the therapy.

N-stage plays a vital role in staging and future treatment process selection since metastasis to the thoracic lymph nodes is the most common way of lung cancer spread. The mediastinal lymph nodes’ involvement in the metastatic process can be a decisive criterion for routing the NSCLC-diagnosed patient either to the radical surgery or the adjuvant therapy Ettinger et al. [2019], Planchard et al. [2018].

Currently, there are two main guidelines for the management of NSCLC-diagnosed patients. The first one proposes performing PET-CT Planchard et al. [2018] and, after that, diagnostic surgical interventions (EBUS/EUS Nakajima et al. [2013], VAMLA Hartert et al. [2020]). The second one says that diagnostic surgery should be conducted, regardless of the PET-CT scanning results Ettinger et al. [2019]. The NCCN guidelines state that surgery should be preferable for patients with early NSCLC, and radiation or chemotherapy in case of advanced NSCLC Ettinger et al. [2017].

However, even using the entire set of tools to detect malignant mediastinal lymph nodes, there is no complete certainty in the lesion or absence of the metastatic process. The misdiagnosis rate Roberts et al. [2000] and the false-negative rate Kanzaki et al. [2011] were higher with PET-CT diagnosis of lymph node metastasis than the results of the histological verification that is considered a golden standard. In addition, PET-CT may not be available for most patients in remote areas Verduzco-Aguirre et al. [2019]. As for the diagnostic surgery, even the minimal invasive requires anesthesia thatmay be contradicted for the patient. Thus, a non-invasive and cost-effective assessment tool is needed to predict the presence of mediastinal lymph node metastases in a patient with primary NSCLC.

Current advances in Deep Learning LeCun et al. [2015] show great potential in solving such problems. Recent studies have reported solid results for both: lymph node stations Guo et al. [2021] and lymph nodes Iuga et al. [2021a,b] segmentation problems. Still, most of these works focus on the algorithmic side of the problem, not the clinical relevance. Moreover, none of them addressed individual lymph node malignancy classification problem, restricting the indirect analysis of the whole study, and limiting the interpretability of the result without giving an option for cliniciansto validate the result Zhong et al. [2018], Liu et al. [2018], Cong et al. [2020].

This work mitigates these gaps, proposing a multi-step algorithm for each visible mediastinal lymph node segmentation and assessing the probability of its involvement in the metastatic process, using the results of histological verification on training. The d*±*eveloped pipeline show 0.74 0.*±*01 average Recall with 0.53 0.26 object Dice Score for the clinically relevant lymph nodes segmentation task and 0.73 ROC AUC for patient’s N-stage prediction, outperforming traditional size-based criteria.

## 2 Related Work

### 2.1 Clinical

Lymph node staging is a vital step for proper NSCLC-verified patient management. To do this, clinicians have a wide spectre of methods, divided into two categories: non-invasive (can be done without intervention to the body) and invasive (assumes diagnostic surgery). The latter show solid results in terms of sensitivity and low post-operative upstaging Gu et al. [2009], Hartert et al. [2020]. However, they require intervention and can be contradicted in some cases because of the anaesthesia. This work will focus on more affordable and widespread non-invasive methods.

Several studies have shown the great limitation of the method, which uses each visible lymph node short-axis diameter (SAD) alone in histological status determination on computed tomography (CT) or magnetic resonance imaging (MRI)scanning. The mesorectal region study reports the vague border between histologically benign (2–10 mm) and malignant (3–15 mm) lymph nodes Brown et al. [2003]. The same problem occurred in the head and neck region. The widely used in clinical practice 10mm cut-off value Som [1987] resulted in sensitivity 0.88 and specificity 0.39 Curtin et al. [1998]. However, the same study shows that other morphological criteria, such as an irregular border or mixed-signalintensity, can improve the results to the sensitivity of 0.85 (95% CI: 0.74, 0.92) and a specificity of 0.97 (95% CI: 0.95, 0.99). A recent study examined the classification accuracy for different combinations of such criteria Loch et al. [2020] but did not propose any standardized way of using them. It was solved with the Node Reporting and Data System (Node-RADS) concept introduction Elsholtz et al. [2021]. The described method hierarchy assesses each visible lymph node, classifying it into five categories, using the SAD, texture, border, and shape criteria, addressing the specifications implied by other factors, such as anatomic location.

A more accurate result can be achieved with the help of contrast-enhanced computed tomography (CE-CT) combined with positron emission tomography (PET). Recent studies show a significant difference (*p <* 0.05) for nodal staging between positron emission tomography combined with computed tomography (PET-CT) with 0.78 sensitivity and 0.92 specificity and 0.56, 0.73 respectively for the CE-CT Ceylan et al. [2012]. Still, PET-CT is a rather expensive method that may not be available for most patients in remote areas Verduzco-Aguirre et al. [2019].

### 2.2 Algorithmic

The mediastinal lymph node segmentation and classification problem has not been studied well by the community, preferably due to the unavailability of good public datasets. Still, there are some works, that address parts of the proposed algorithm in this paper.

#### Lymph Nodes Segmentation

Over the past years many approaches were introduced to solve the volumetric medical images segmentation problem. Architectures, such as DeepMedic Kamnitsas et al. [2017], 3D U-Net Çiçek et al. [2016] or V-Net Milletari et al. [2016] provide solid results on public datasets with medical images Van Ginneken et al. [2010], Bakas et al. [2018]. The proposed feature-pyramidal (FPN) convolutional neural networks (CNN) were successfully adapted to the lymph node segmentation problem. In the most relevant work Iuga et al. [2021a], the authors report a total detection rate of 0.77 for enlarged lymph nodes during fourfold cross-validation with 10.3 false positives (FP) per case. However, showing poor sensitivity the lymph nodes of diameter from 5mm to 10mm 0.34 and a rather low overall Dice Score (DSC) of 0.44.

#### Lymph Node Stations Identification

The paper Iuga et al. [2021b] that deals with the distribution of the lymph nodes to the mediastinal stations utilizes the same architecture as Iuga et al. [2021a], but with the multiclass output in the end. The authors report convincing mean classification accuracies: 0.86 (Top-1), 0.94 (Top-2) and 0.96 (Top-3). However, the proposed algorithm still has poor sensitivity for the most important lymph node stations, also only indirectly addressing the guidelines Goldstraw et al. [2016]. More advanced work solves the segmentation task Guo et al. [2021] reporting DSC 0.81*±*0.06, but skipping the accuracy of the lymph nodes distribution and its influence on the N-stage determination.

#### Lymph Nodes Malignancy Classification

Previous works proposed algorithms for the indirect analysis of mediastinal lymph nodes’ involvement in the metastatic process. The popular approach uses a set of radiomic features, extracted from a primary tumor Zhong et al. [2018], Liu et al. [2018], Cong et al. [2020], without specifying the particular lymph node station, not saying about the concrete node. One of the reasons for that is the complexity of obtaining the ground truth label for each lymph node. After the biomaterial extraction, it is hard to say the position of each lymph node on the CT scan, especially for the small ones. The solution would be straightforward, having a label for each lymph node, as was shown in the similar problem of lung nodules malignancy classification. Simple patch-based convolutional neural network for classification results in ROC AUC of 0.928 0.027. Better quality can be achieved via pre-training the convolutional autoencoder for the extracted patch reconstruction and utilization of the encoder as a backbone for malignancy classification, giving ROC AUC of 0.936 0.009 Silva et al. [2020]. However, it is possible to get the general histological label for a group of lymph nodes, extracted from the same station, formulatingthe problem as a weak-supervision task. An interesting idea was proposed in Dubost et al. [2020] by using the single label for training while predicting the probability map on inference. However, the global max-pooling (GMP) operation will lose too much information for such small objects as lymph nodes, unlikely to be successful for the histological status determination problem.

## 3 Method

The proposed algorithm for lymph node segmentation and malignancy classification is three-step. First, the mediastinumis segmented to narrow down the region of interest and identify lymph node stations that play a vital role in the N-stagedetermination Detterbeck et al. [2017]. Then, the mediastinum region’s bounding box is used to crop the input imageand segment each visible lymph node. Finally, each of the detected lymph nodes is passed through the feed-forward network to get its probability of being malignant. The generated result contains information about malignant lymph nodes found in a particular lymph node station, and, depending on the tumor side – the desired N-stage. Each part ofthe developed pipeline is described in a specific section:

- Subsection 3.1 is devoted to the lymph node stations segmentation;
- Subsection 3.2 is devoted to the lymph nodes segmentation;
- Subsection 3.3 is devoted to the lymph nodes malignancy classification;

### 2.3 Lymph Node Stations Segmentation

The lymph nodes involved in the N-staging of NSCLC-diagnosed patients are located in a narrow region – mediastinum. Moreover, N-stage determination depends on the particular anatomic location (called “station”) and the primary tumorside Detterbeck et al. [2017]. According to the IASLC Rusch et al. [2009] the mediastinum is divided into 10 lymph node stations. Adjacent to the trachea and bronchi stations are subdivided by the left and right side, except the subcarinal, which has no specific subdivision. The diagnostic surgeries usually don’t take biomaterial from stations 1, 8, and 9, sothis work waves them out.

This work utilizes de-facto 3D implementation of U-Net Ronneberger et al. [2015] with two heads for the stations’ segmentation (Figure 1). The first separates the mediastinum from the background. The second classifies each voxel inside the mediastinal mask to one of the lymph node stations. Moreover, we use all the best practices introduced by the deep learning community over the recent years: residual blocks (ResBlocks) He et al. [2016], batch normalization Ioffe and Szegedy [2015] and ReLU activation function Nair and Hinton [2010] after every convolution except the output.

**Figure 1:**
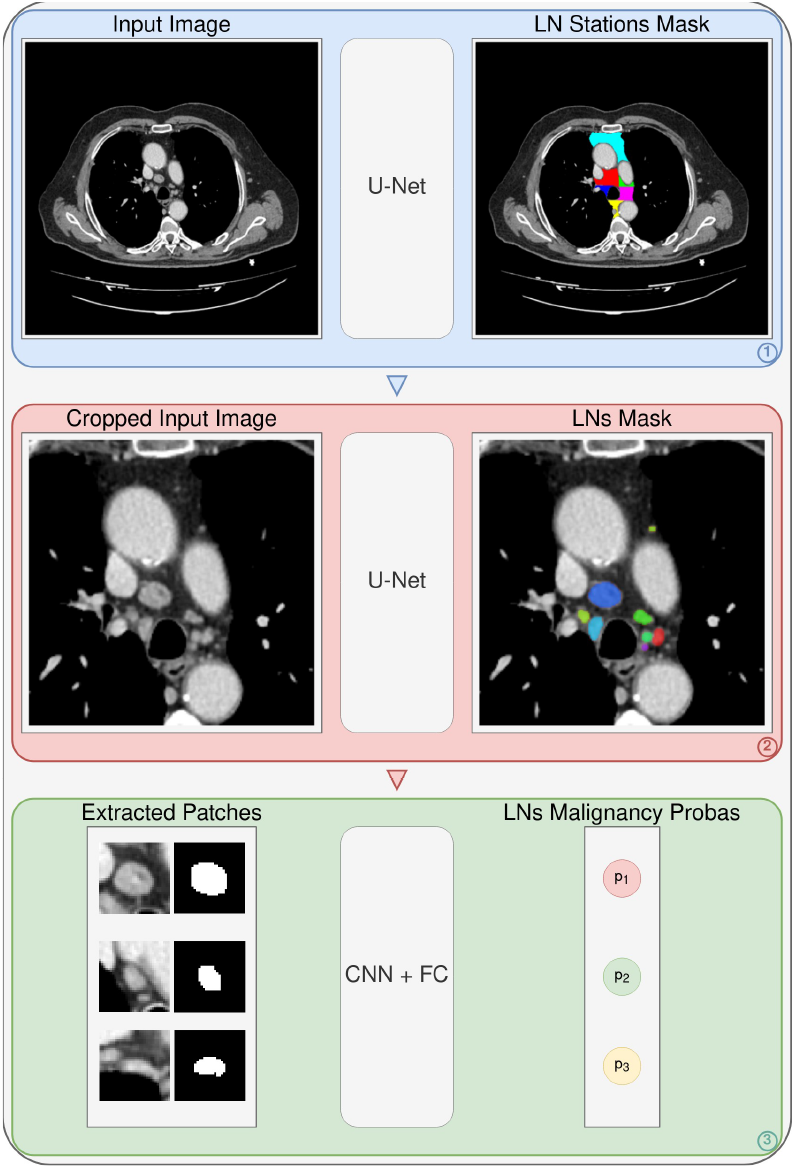
The proposed multi-step pipeline for lymph nodes segmentation and malignancy classification. In the first step, the IASLC lymph node stations are segmented, also giving the bounding box of the mediastinum for the next step. Then, the image is cropped to that box and passed through the second network to get the mask of all visible lymph nodes. Finally, each detected lymph node is extracted from the image, stacked with its mask and processed through the feed-forward network to get the corresponding malignancy probabilities.

**Figure 2:**
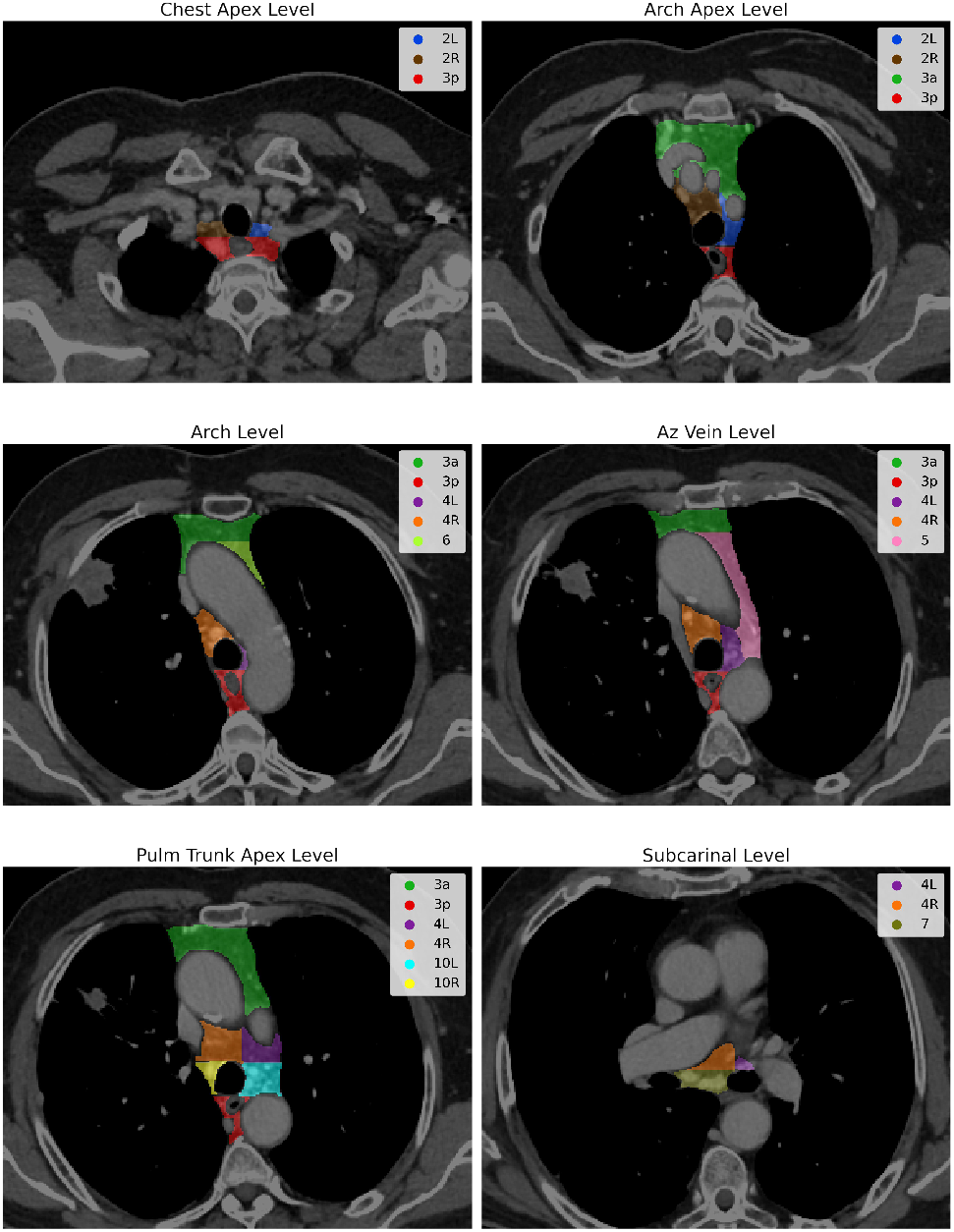
The example of the lymph node stations annotation at different mediastinum levels.

### 2.4 Lymph Nodes Segmentation

Despite the image itself, the second step requires a mediastinum box from the first step to extract the narrow region for the computational resources economy. If the box size is less than 128 pixels in the Axial projection, it is additionally padded to the minimal shape of 128 pixels.

In the end, the architecture of the second step remains practically the same as for lymph node station segmentation, except having fewer levels and more channels with the single binary output for the predicted lymph nodes segmentation map (Figure 1). This design solution is motivated by the fact that lymph nodes are much smaller objects than the stations and do not require a large receptive field, but the additional features are helpful for their segmentation.

### 2.5 Lymph Nodes Malignancy Classification

The task is formulated as a weak supervision problem: having a malignancy status label for the whole station, and predicting the probabilities of metastases for each of the included lymph nodes. To do this, the patches of a fixed size 32 *×*32 with lymph nodes are extracted from the image and combined with the corresponding mask, gathering all the detected objects in a single batch. Then, it is processed through the ResNet-like He et al. [2016] convolutional neural network (CNN) of 5 levels, followed by a max-pooling layer, shrinking the spatial dimensions, and, finally, passed through the fully connected and sigmoid layers to get the final probabilities for each lymph node to be malignant.

The additional complexity is that the malignant station, in contrast to the benign, can contain as benign as malignant lymph nodes. Thus, it can be considered that a benign station has no malignant lymph nodes, but least the malignant station must have at least one. A specific loss function is developed to address this rule. All the lymph node probabilities in the benign station are trained with binary cross-entropy (BCE) loss, but, the malignant nodes – only if every node in that zone was predicted as benign. This method has its upsides and downsides that are thoroughly discussed in Section5.

## 4 Experiment

### 4.1 Data

Unfortunately, the only available public dataset Roth et al. [2014] has several limitations. First, there is not enough information about the diagnosis and histological status of mediastinal lymph nodes. Second, the provided annotation contains LN only with a short-axis diameter (SAD) greater than 10mm and no lymph node stations annotation. Finally, the contrast phase of the contrast-enhanced computed tomography (CE-CT) scans seems to be arterial instead of venous. So, the private dataset of 60 patients who had verified NSCLC and had undergone diagnostic surgery to examine someof the lymph node stations was acquired.

The following inclusion criteria were applied to the collected dataset:

- presence of venous contrast phase, because it provides the best differentiation of lymph nodes from the surrounding structures, especially vessels;
- diagnostic surgery was conducted no later than two months from the latest study, which included the venous phase;
- slice thickness of CE-CT should not be greater than 1mm; what resulted in 8 series that we gave for annotation.

#### 4.1.1 Annotation

##### Lymph Node Stations

The lymph node stations annotation was done by a single radiologist, in strict accordance with the IASLC guidelines for mediastinum map Rusch et al. [2009]. The annotation protocol was ordered to exclude the big vessels (aorta, pulmonary trunk, Azygos vein, etc.) and oesophagus from the station areas 2.

##### Lymph Nodes

The mediastinal lymph node annotation has been conducted by two radiologists, who have assigned each visible lymph node its binary mask. In the case of multiple lymph nodes having an indistinct border between themselves, a single mask was assigned to the whole conglomerate.

##### Lymph Node Staging

The mediastinal lymph node station malignancy was obtained after conducting VAMLA Hartert et al. [2020] diagnostic surgery. The extracted nodes were passed to the biopsy, where the status of each extracted lymph node was determined. Finally, each station was assigned one of three labels, depending on the histological analysis result: “n/a” (because the station was not resected), “benign” or “malignant”. The training dataset statistics canbe found in Table 1.

**Table 1:** The training dataset biopsy result statistics for IASLC lymph node stations.

| Status | 1R | 1L | 2R | 2L | 3a | 3p | 4R | 4L | 5 | 6 | 7 | 8 | 9 | 10R | 10L |
| --- | --- | --- | --- | --- | --- | --- | --- | --- | --- | --- | --- | --- | --- | --- | --- |
| benign | 0 | 0 | 5 | 3 | 0 | 0 | 6 | 8 | 0 | 0 | 6 | 0 | 1 | 2 | 2 |
| malignant | 0 | 0 | 2 | 0 | 0 | 0 | 2 | 0 | 1 | 1 | 1 | 0 | 0 | 2 | 1 |
| n/a | 8 | 8 | 1 | 5 | 8 | 8 | 0 | 0 | 7 | 7 | 1 | 8 | 7 | 4 | 5 |

**Table 2:**
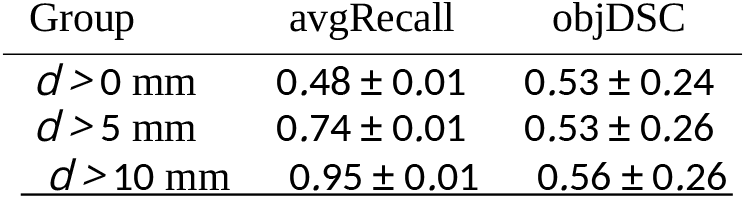
Lymph nodes detection metrics for different SAD groups.

| Group | avgRecall | objDSC |
| --- | --- | --- |
| $d > 0$ mm | $0.48 \pm 0.01$ | $0.53 \pm 0.24$ |
| $d > 5$ mm | $0.74 \pm 0.01$ | $0.53 \pm 0.26$ |
| $d > 10$ mm | $0.95 \pm 0.01$ | $0.56 \pm 0.26$ |

**Table 3:** Patients’ N-status, the maximum SAD and predicted malignancy probability. Note, that lymph nodes chosen by the diameter criteria can differ from the ones with maximum malignancy probability.

| Patient | N | max $d$, mm | max $p$ |
| --- | --- | --- | --- |
| 0 | 0 | 11 | 0.29 |
| 1 | 0 | 14 | 0.41 |
| 2 | + | 38 | 1.00 |
| 3 | + | 14 | 0.54 |
| 4 | + | 14 | 0.54 |
| 5 | 0 | 21 | 0.83 |
| 6 | + | 15 | 0.61 |
| 7 | + | 11 | 0.96 |

### 4.2 Training

In all experiments was utilized the standard preprocessing, which zooms the input image to the constant voxel spacing of (1, 1, 1), clips the input CT scan to the soft-tissue window of [ 160; 240] HU, and, finally, scale the intensities to the [0; 1]. The only difference is that for the first step, the input image is cropped to the lungs, using the pre-trained neural network Goncharov et al. [2021] and the second step is cropped to the mediastinum. Also, being in a situation with a tiny dataset, severe augmentations are applied during training: rotation for the limit of 10 degrees, 90-degree rotation,random shifts, and vertical and horizontal flips.

#### Lymph Node Stations Segmentation

The first step is trained for 30k iterations of Adam, using mixed-precision and gradient scaling. For the first head, binary cross-entropy (BCE) with adaptive re-weighting of the foreground voxels is utilized, and, for the second – cross-entropy (CE). The learning rate remains the same at the level of 3 10^−3^ during the training process. *·*

#### Lymph Nodes Segmentation

The second step is trained for 70k iterations of Adam, using mixed-precision and gradient scaling, minimizing BCE, and adaptively re-weighting the foreground voxels, the same way as in the first step. The training starts with a learning rate of 10^−3^ and is being reduced by a factor of 3, 3, 2, and 2 at the 5k, 15k, 50k, and 60k iterations respectively.

#### Lymph Node Malignancy Classification

The classification network is trained for 40k iterations of Adam, minimizing the loss, described in Subsection 3.3, re-weighting the positive-case examples by a factor of 100. The training starts witha learning rate of 3 *·* 10^−5^ and remains the same till the end.

### 4.3 Metrics

In assessing the first step lymph node assignment accuracy to the particular station is measured, instead of the classical segmentation metrics, because they are less informative in this particular case. Also, for the third step, prediction can only be assessed by the final result, where the determined N-stage ROC AUC score is computed for N-positive and N-negative patients. Metrics for the second step are described in more detail below.

#### 4.3.1 FROC

To assess the lymph nodes detection quality we use the Free-response Receiver Operating Characteristic (FROC) analysis Van Ginneken et al. [2010]. Such a curve illustrates the trade-off between the model’s object-wise recall (Y-axis) and the average number of false positives (FPs) per image (X-axis).

To build one, we take the ground truth mask of the whole image (Figure 3 a) and the corresponding map of logits, binarized using the threshold value of 0 to obtain logit mask (Figure 3 b). Then, we split both masks into the connected components (CCs) and to each CC we assign 3 statistics:

**Figure 3:**
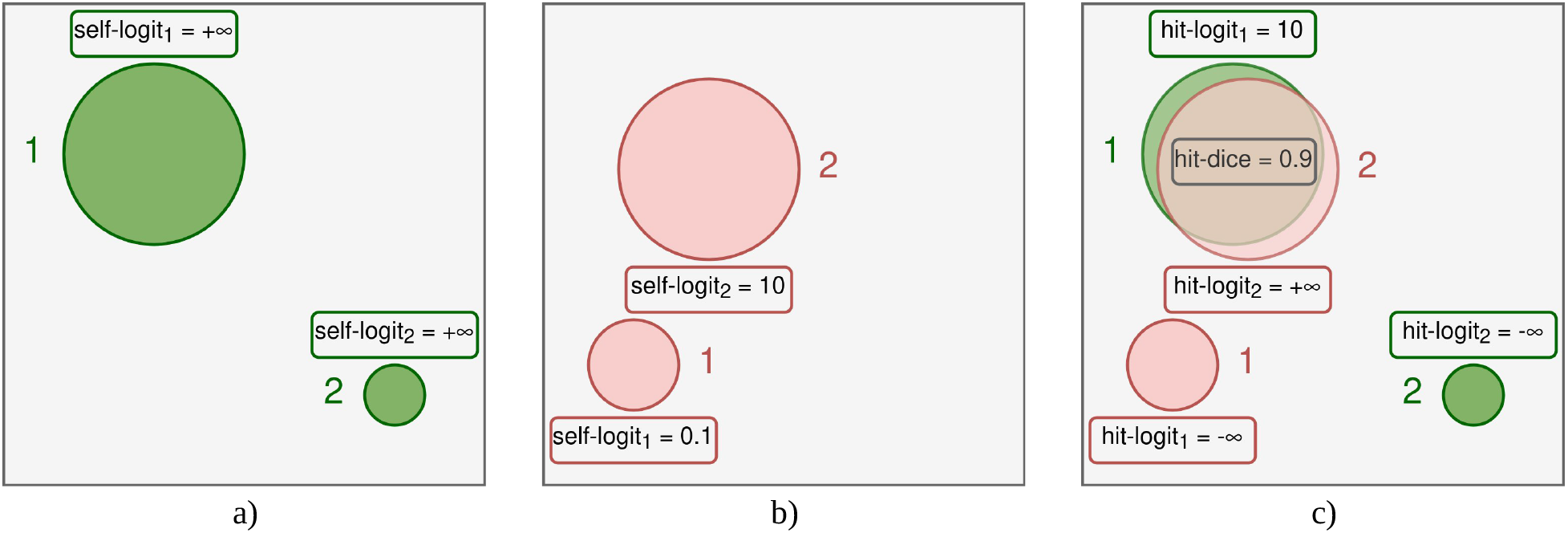
Illustration to the way considered statistics are assigned to different connected components (CCs) inside the ground truth mask (Figure a) and the corresponding logit mask (Figure b). Although *self-logit* and *hit-logit* are a kind of personal statistics for each CC, *hit-dice*, as can be seen from the (Figure c), is always shared between the pair of CCs if its value is positive.

**Figure 4:**
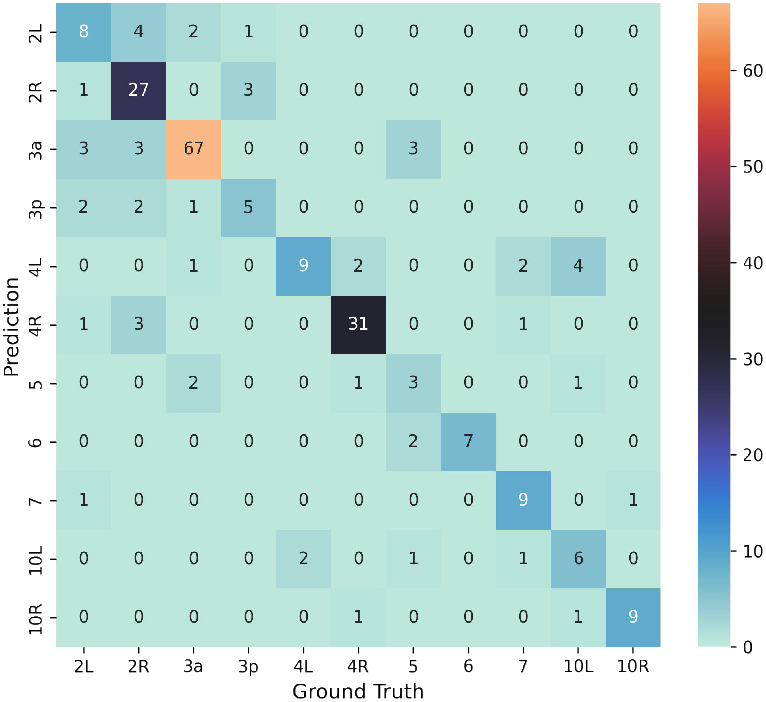
Accuracy of the IASLC lymph node stations assignment.

- *self-logit* — the maximum logit value inside the CC (+∞ if the CC is located inside the ground truth mask),
- *hit-dice* — the maximum Dice Score between the chosen CC and the CC from the other mask (Figure 3 c),
- *hit-logit* — the same statistics as *self-logit*, but taken from the CC inside the other mask that hits the first one with the *hit-dice* value (we consider it as −∞ if *hit-dice* is 0).

Based on these statistics we can take *hit-logit* as the value we will use to build the FROC curve, choosing different *l-threshold* values. Moreover, we also take *hit-dice* into account while obtaining the points of the curve, to check the so called *hit condition*. We say that *hit condition* between the two CCs is satisfied if *hit-dice* value is positive. So that, for the chosen *l-threshold* value we define:

- **FP** value as the number of CCs inside the logit-mask that has

*self-logit > l-threshold*, but *hit-dice* = 0;

- **TP** value as the number of CCs inside the ground-truth mask that has

*hit-logit > l-threshold* and *hit-dice* > 0;

- **FN** value as the number of CCs inside the ground-truth mask that has

*hit-logit ≤ l-threshold* or *hit-dice* = 0.

In our experiments, we choose *l-threshold* values from the ( 0.1; *max-logit*) range, where *max-logit* is the maximum logit value over the *hold-out* set of predictions. −

In contrast to the classical approach of building such curves, the chosen method guarantees the monotony. This comes from the property of any CC inside the prediction mask. It appears in the mask fully or doesn’t appear at all instead of continuous change in the classical approach, that may lead to confusing situations like splitting a single CC into many distinct CCs or vice versa. The choice to iterate over the logit values instead of classical probabilities is motivated by the floating-point precision restrictions. This scale allows us to build full-ranged curves because large logit values can be separated with much higher precision than the large probabilities, which tend to round upward to 1.

#### 4.3.2 Average Recall

Although FROC analysis provides a very deep understanding of the obtained results, it still can be hard to understand. For this reason, the next metric is chosen as the simplification and generalization of information that FROC curves give. In our work, we report values averaged over FP points from 0 to 5 with a step of 0.01. Hence, the detection quality ismeasured similarly. This is our main quality metric because the fraction of detected lesions per case is an important clinical characteristic.

#### 4.3.3 Object Dice Score

The most common method to measure segmentation quality is the Dice Score Bakas et al. [2018]. However, averaging the Dice Score (DSC) over images has a serious drawback in the case of multiple targets because large objects overshadow small ones. Hence, we report the average *object-wise* Dice Score:

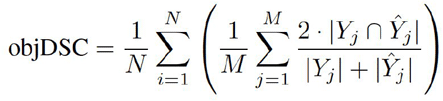

where *N* is the number of images in the *hold-out* set, *M* is the number of lesions inside the ground truth mask, *Y*_*j*_ — the set of voxels, that relates to the *j*-th CC inside that mask and *Ŷ*_*j*_ the corresponding CC from the prediction mask that has the biggest overlap with it in terms of Dice Score.

## 5 Results

### Lymph Nodes Segmentation

The Second step’s metric is reported for different size ranges: “all lymph nodes”, “greater than 5 mm” (treated as clinically relevant by to the guidelines), and “greater than 10 mm” (used in the simplest criteria for a node to be metastasized) Elsholtz et al. [2021]. Despite the poor detection rate in the first group, the CNN shows a perfect sensitivity for the riskiest group (the last one) at the level of only three false positives (FP) per case (Figure 5).

**Figure 5:**
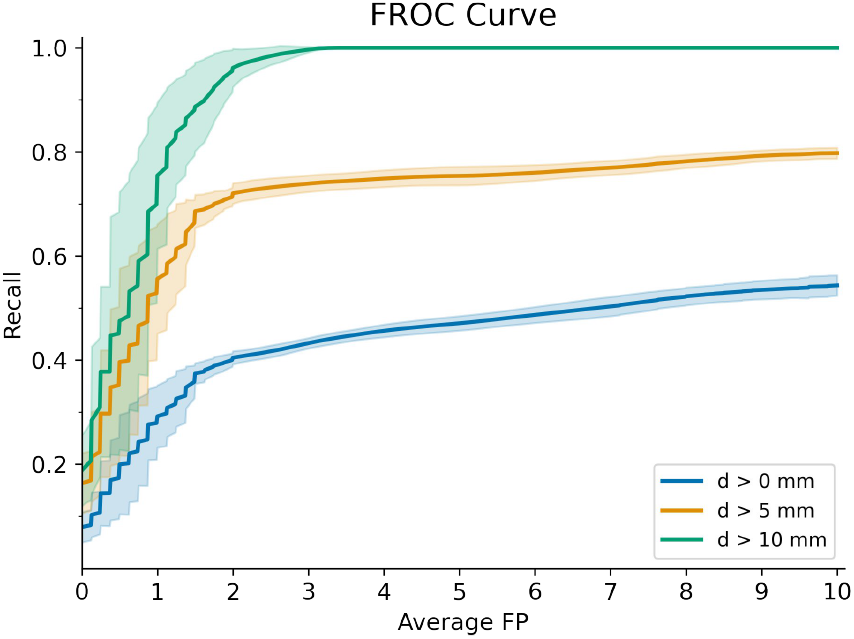
Detection results for the lymph nodes segmentation step. The 80% of clinically relevant nodes are successfully detected by the algorithm at the level of 10 FP per case. Moreover, the network doesn’t miss any of the large lymph nodes with only 3 FP per case.

### Lymph Node Malignancy Classification

The obtained malignancy classification results were compared to a naive approach, where the lymph node with maximal short-axis diameter (SAD) was taken. This simple criterion yields three FP results (the 10mm threshold), however, the proposed algorithm outperforms it (Figure 6) showing a better ROC AUC score of 0.73 in comparison to the 0.53 for a naive approach. Note, that following the Node-RADS Elsholtz et al. [2021] guidelines, the larger than 30 mm SAD lymph nodes are treated as malignant for sure. The only mistake was made for patient 5, where the highlighted lymph node is very suspicious of metastatic involvement.

**Figure 6:**
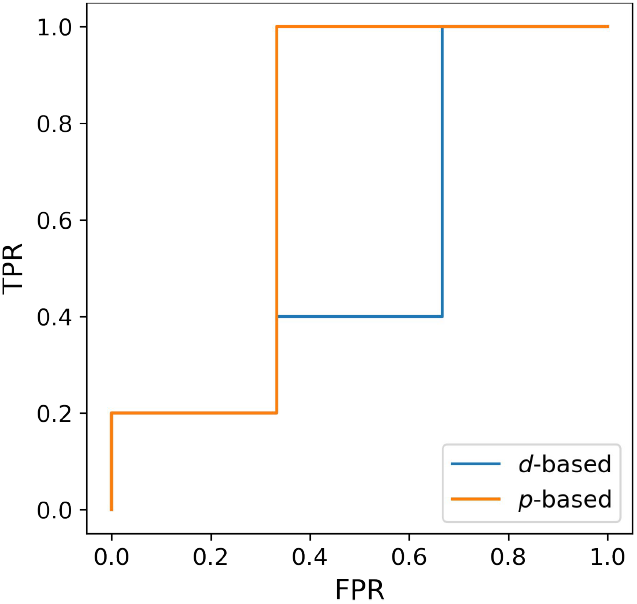
Comparison between the naive SAD-based criteria for patient’s N-status prediction with the proposedalgorithm.

**Figure 7:**
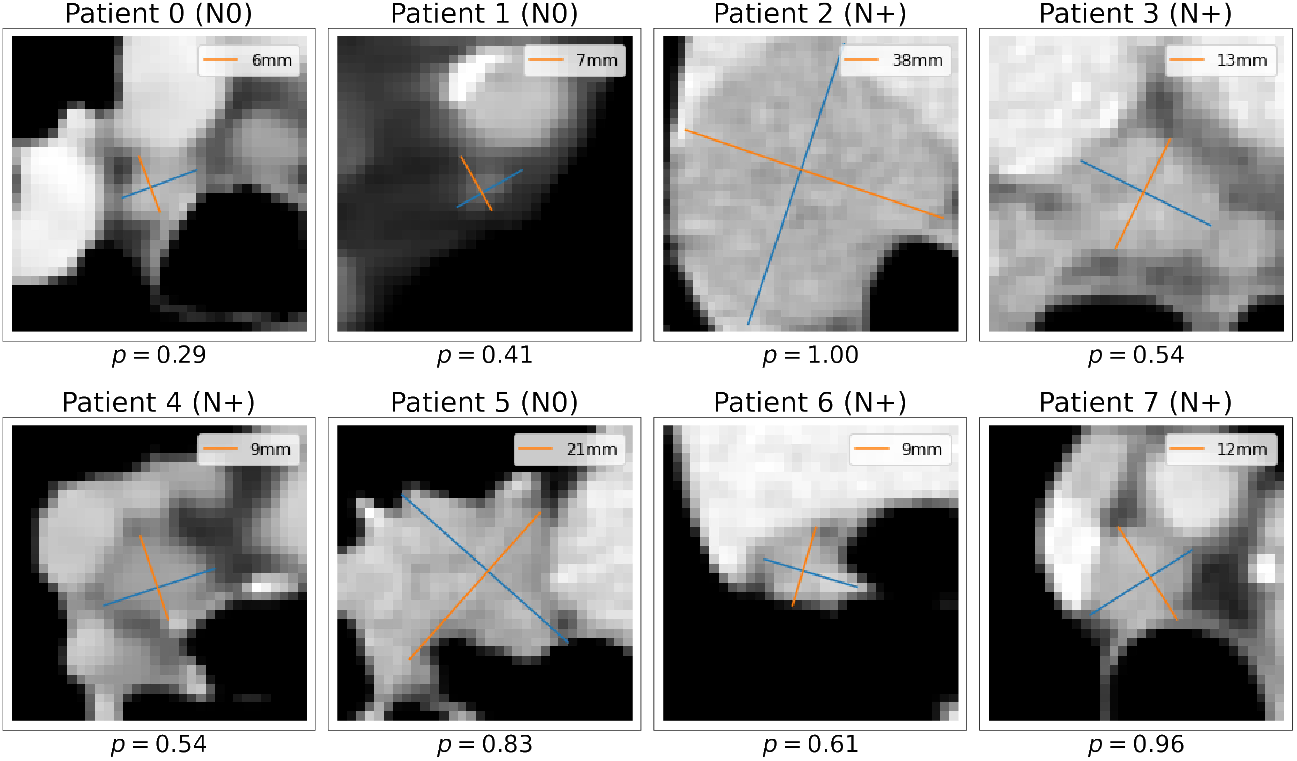
Lymph nodes with the maximum malignancy probability for each patient.

## 6 Discussion

The proposed loss function has its upsides and downsides. On the one hand, the CNN is free to decide the relation between lymph nodes assigned to the malignant station and based on its negative experience balance it. On the other, there is no reinforcement for assigning the right probabilities for positive-class examples, because there is no information for each lymph node in the malignant station. This way of learning can result in higher sensitivity but produce unpleasant FPs.

Since this task places a high level of demand on the localization of lymph nodes based on clinical classification, the main limitation of this work was the small size of the dataset, providing an insufficient variety of examples of benign and malignant lymph nodes in each group. The major reason is a very tedious and time-consuming process of lymph nodestations and lymph node delineation from scratch. The lymph nodes, it is given for two to three hours for one patient, and the lymph node stations – about 1 hour, with very vague criteria of human anatomy, making it very difficult to derive general rules. In this regard, it is expected to improve the performance indicators of the algorithm when expanding thetraining set, primarily by including new cases with non-enlarged metastatic nodes and enlarged non-metastatic ones for each group of intrathoracic lymph nodes.

Important information to help solve the problem can be taken from data using multiphase CT scans so that information is available for each lymph node without and with intravenous contrast. First of all, the use of the venous phase of intravenous contrast has been demonstrated in similar works as the most valuable CT series Cong et al. [2020]. However, the generalized value of information from multiphase scanning, including native (non-contrast), arterial, venous and delayed phases of intravenous contrast, remains unexplored, which may provide additional information about the accumulation and washout of contrast agent from each lymph node.

## 7 Conclusion

This work proposes a three-step pipeline for the lymph node segmentation and malignancy classification for NSCLC-diagnosed patients, using histological verification information about lymph node stations on training. The developed pipeline shows 0.74*±*0.01 average Recall with 0.53 *±*0.26 object Dice Score for the clinically relevant lymph nodes (SAD ≥ 5mm) segmentation task and 0.73 ROC AUC for patient’s N-stage prediction, outperforming traditional size-based criteria. The segmentation performance increases up to 0.95 *±* 0. 01 average Recall with 0.56 *±* 0.26 object Dice Score for the enlarged lymph nodes (SAD ≥ 10mm) and makes it possible to conduct new research to optimize the management of patients with non-enlarged intrathoracic lymph nodes, improving the quality of medical care for cancer patients.

In perspective, the algorithm proposed in this paper can be integrated into the existing routing of patients with verified NSCLC as an intermediate step between the confirmation of the primary diagnosis and PET-CT scanning. There are multiple variants of the algorithm that can be useful. In the first scenario, where the algorithm will give a low probability of having a metastatic lesion of the lymph nodes of the mediastinum, it will allow the patient to be sent for radical surgery, skipping the steps of PET-CT, as well as performing a diagnostic surgery. In the case of a high probability, the patient will immediately be contraindicated for radical surgery and given a referral for neoadjuvant chemotherapy, again bypassing the steps of PET-CT and diagnostic surgery. Even operating at a level of accuracy comparable to PET-CT,the algorithm will be significantly cheaper to use and also much more accessible to patients.

## Data Availability

All data produced in the present study are available upon reasonable request to the authors.

## Acknowledgments

The authors would like to thank Shukran Ragimov, and Anatolii Akhmedov for the IASLC lymphnode stations biopsy results extraction, and Anastasia Nikulina and Ekaterina Chukanova for lymph node annotation.

## Notes

### Competing Interest Statement

The authors have declared no competing interest.

### Funding Statement

This study did not receive any funding.

### Author Declarations

The Independent Ethical Committee of the Moscow Regional Branch of the Russian Society of Radiologists gave ethical approval for this work dated 23 March 2023, protocol no. 03/2023

## References

Krishna Chaitanya Thandra, Adam Barsouk, Kalyan Saginala, John Sukumar Aluru, and Alexander Barsouk. Epidemiology of lung cancer. Contemporary Oncology, 25(1):45, 2021.

Peter Goldstraw, Kari Chansky, John Crowley, Ramon Rami-Porta, Hisao Asamura, Wilfried EE Eberhardt, Andrew G Nicholson, Patti Groome, Alan Mitchell, Vanessa Bolejack, et al. The iaslc lung cancer staging project: proposals for revision of the tnm stage groupings in the forthcoming (eighth) edition of the tnm classification for lung cancer. Journal of Thoracic Oncology, 11(1):39–51, 2016.

Lynn T Tanoue, Nichole T Tanner, Michael K Gould, and Gerard A Silvestri. Lung cancer screening. American journal of respiratory and critical care medicine, 191(1):19–33, 2015.

David S Ettinger, Douglas E Wood, Charu Aggarwal, Dara L Aisner, Wallace Akerley, Jessica R Bauman, Ankit Bharat, Debora S Bruno, Joe Y Chang, Lucian R Chirieac, et al. Nccn guidelines insights: non–small cell lung cancer, version 1.2020: featured updates to the nccn guidelines. Journal of the National Comprehensive Cancer Network, 17(12):1464–1472, 2019.

D Planchard, ST Popat, K Kerr, S Novello, EF Smit, Corinne Faivre-Finn, TS Mok, M Reck, PE Van Schil, MD Hellmann, et al. Metastatic non-small cell lung cancer: Esmo clinical practice guidelines for diagnosis, treatment and follow-up. Annals of Oncology, 29:iv192–iv237, 2018.

Bruno Heleno, Volkert Siersma, and John Brodersen. Estimation of overdiagnosis of lung cancer in low-dose computed tomography screening: a secondary analysis of the danish lung cancer screening trial. JAMA internal medicine, 178(10):1420–1422, 2018.

Andrea Lopes Pegna, Giulia Picozzi, Fabio Falaschi, Laura Carrozzi, Massimo Falchini, Francesca Maria Carozzi, Francesco Pistelli, Camilla Comin, Annalisa Deliperi, Michela Grazzini, et al. Four-year results of low-dose ct screening and nodule management in the italung trial. Journal of Thoracic Oncology, 8(7):866–875, 2013.

Maurizio Infante, Silvio Cavuto, Fabio Romano Lutman, Eliseo Passera, Maurizio Chiarenza, Giuseppe Chiesa, Giorgio Brambilla, Enzo Angeli, Giuseppe Aranzulla, Arturo Chiti, et al. Long-term follow-up results of the dante trial, a randomized study of lung cancer screening with spiral computed tomography. American journal of respiratory andcritical care medicine, 191(10):1166–1175, 2015.

H de Koning, C van der Aalst, P de Jong, et al. Screening met een thoracale lage-dosis-ct-scan vermindert de sterfte na 10 jaar door longkanker bij mannelijke actieve of ex-rokers. N. Engl. J. Med, 382:503–513, 2020.

U Pastorino, M Silva, S Sestini, F Sabia, M Boeri, A Cantarutti, N Sverzellati, G Sozzi, G Corrao, and A Marchianò. Prolonged lung cancer screening reduced 10-year mortality in the mild trial: new confirmation of lung cancer screening efficacy. Annals of Oncology, 30(7):1162–1169, 2019.

DR Baldwin, SW Duffy, NJ Wald, R Page, DM Hansell, and JK3063456 Field. Uk lung screen (ukls) nodule management protocol: modelling of a single screen randomised controlled trial of low-dose ct screening for lung cancer. Thorax, 66(4):308–313, 2011.

Frank C Detterbeck, Daniel J Boffa, Anthony W Kim, and Lynn T Tanoue. The eighth edition lung cancer stage classification. Chest, 151(1):193–203, 2017.

Takahiro Nakajima, Kazuhiro Yasufuku, and Ichiro Yoshino. Current status and perspective of ebus-tbna. General thoracic and cardiovascular surgery, 61(7):390–396, 2013.

Marc Hartert, Jan Tripsky, and Martin Huertgen. Video-assisted mediastinoscopic lymphadenectomy (vamla) for staging & treatment of non-small cell lung cancer (nsclc). Mediastinum, 4, 2020.

David S Ettinger, Douglas E Wood, Dara L Aisner, Wallace Akerley, Jessica Bauman, Lucian R Chirieac, Thomas A D’Amico, Malcolm M DeCamp, Thomas J Dilling, Michael Dobelbower, et al. Non–small cell lung cancer, version 5.2017, nccn clinical practice guidelines in oncology. Journal of the National Comprehensive Cancer Network, 15 (4):504–535, 2017.

Peter F Roberts, David M Follette, Derek von Haag, Jason A Park, Peter E Valk, Thomas R Pounds, and Donald M Hopkins. Factors associated with false-positive staging of lung cancer by positron emission tomography. The Annalsof thoracic surgery, 70(4):1154–1159, 2000.

Ryu Kanzaki, Masahiko Higashiyama, Ayako Fujiwara, Toshiteru Tokunaga, Jun Maeda, Jiro Okami, Takenori Kozuka, Takuya Hosoki, Yoshihisa Hasegawa, Motohisa Takami, et al. Occult mediastinal lymph node metastasis in nsclc patients diagnosed as clinical n0-1 by preoperative integrated fdg-pet/ct and ct: risk factors, pattern, and histopathological study. Lung cancer, 71(3):333–337, 2011.

Haydeé C Verduzco-Aguirre, Gilberto Lopes, and Enrique Soto-Perez-De-Celis. Implementation of diagnostic resources for cancer in developing countries: a focus on pet/ct. ecancermedicalscience, 13, 2019.

Yann LeCun, Yoshua Bengio, and Geoffrey Hinton. Deep learning. nature, 521(7553):436–444, 2015.

Dazhou Guo, Xianghua Ye, Jia Ge, Xing Di, L. Lu, Lingyun Huang, Guotong Xie, Jing Xiao, Zhongjie Lu, Ling Peng,et al. Deepstationing: thoracic lymph node station parsing in ct scans using anatomical context encoding and key organ auto-search. In International Conference on Medical Image Computing and Computer-Assisted Intervention, pages 3–12. Springer, 2021.

Andra-Iza Iuga, Heike Carolus, Anna J Höink, Tom Brosch, Tobias Klinder, David Maintz, Thorsten Persigehl, Bettina Baeßler, and Michael Püsken. Automated detection and segmentation of thoracic lymph nodes from ct using 3d foveal fully convolutional neural networks. BMC Medical Imaging, 21(1):1–12, 2021a.

Andra-Iza Iuga, Tanja Lossau, Liliana Laurenco Caldeira, Miriam Rinneburger, Simon Lennartz, Nils Große Hokamp, Michael Püsken, Heike Carolus, David Maintz, Tobias Klinder, et al. Automated mapping and n-staging of thoracic lymph nodes in contrast-enhanced ct scans of the chest using a fully convolutional neural network. European Journalof Radiology, 139:109718, 2021b.

Yan Zhong, Mei Yuan, Teng Zhang, Yu-Dong Zhang, Hai Li, and Tong-Fu Yu. Radiomics approach to prediction of occult mediastinal lymph node metastasis of lung adenocarcinoma. American Journal of Roentgenology, 211(1): 109–113, 2018.

Ying Liu, Jongphil Kim, Yoganand Balagurunathan, Samuel Hawkins, Olya Stringfield, Matthew B Schabath, Qian Li, Fangyuan Qu, Shichang Liu, Alberto L Garcia, et al. Prediction of pathological nodal involvement by ct-based radiomic features of the primary tumor in patients with clinically node-negative peripheral lung adenocarcinomas. Medical physics, 45(6):2518–2526, 2018.

Mengdi Cong, Haoyue Yao, Hui Liu, Liqiang Huang, and Gaofeng Shi. Development and evaluation of a venous computed tomography radiomics model to predict lymph node metastasis from non-small cell lung cancer. Medicine, 99(18), 2020.

Ping Gu, Yi-Zhuo Zhao, Li-Yan Jiang, Wei Zhang, Yu Xin, and Bao-Hui Han. Endobronchial ultrasound-guided transbronchial needle aspiration for staging of lung cancer: a systematic review and meta-analysis. European journalof cancer, 45(8):1389–1396, 2009.

Gina Brown, Catherine J Richards, Michael W Bourne, Robert G Newcombe, Andrew G Radcliffe, Nicholas S Dallimore, and Geraint T Williams. Morphologic predictors of lymph node status in rectal cancer with use of high-spatial-resolution mr imaging with histopathologic comparison. Radiology, 227(2):371–377, 2003.

Peter M Som. Lymph nodes of the neck. Radiology, 165(3):593–600, 1987.

Hugh D Curtin, Hemant Ishwaran, Anthony A Mancuso, Robert W Dalley, Daryl J Caudry, and Barbara J McNeil. Comparison of ct and mr imaging in staging of neck metastases. Radiology, 207(1):123–130, 1998.

Florian N Loch, Patrick Asbach, Matthias Haas, Hendrik Seeliger, Katharina Beyer, Christian Schineis, Claudius E Degro, Georgios A Margonis, Martin E Kreis, and Carsten Kamphues. Accuracy of various criteria for lymph node staging in ductal adenocarcinoma of the pancreatic head by computed tomography and magnetic resonance imaging. World Journal of Surgical Oncology, 18(1):1–10, 2020.

Fabian HJ Elsholtz, Patrick Asbach, Matthias Haas, Minerva Becker, Regina GH Beets-Tan, Harriet C Thoeny, Anwar R Padhani, and Bernd Hamm. Introducing the node reporting and data system 1.0 (node-rads): a concept for standardized assessment of lymph nodes in cancer. European radiology, 31(8):6116–6124, 2021.

Naim Ceylan, Sozen Dogan, Kenan Kocaçelebi, Recep Savaş, Alpaslan Çakan, and Ufuk Cagrici. Contrast enhanced ct versus integrated pet-ct in pre-operative nodal staging of non-small cell lung cancer. Diagnostic and InterventionalRadiology, 18(5), 2012.

Konstantinos Kamnitsas, Christian Ledig, Virginia FJ Newcombe, Joanna P Simpson, Andrew D Kane, David K Menon, Daniel Rueckert, and Ben Glocker. Efficient multi-scale 3d cnn with fully connected crf for accurate brain lesion segmentation. Medical Image Analysis, 36:61–78, 2017.

Özgün Çiçek, Ahmed Abdulkadir, Soeren S Lienkamp, Thomas Brox, and Olaf Ronneberger. 3d u-net: learning dense volumetric segmentation from sparse annotation. In International conference on medical image computing and computer-assisted intervention, pages 424–432. Springer, 2016.

Fausto Milletari, Nassir Navab, and Seyed-Ahmad Ahmadi. V-net: Fully convolutional neural networks for volumetric medical image segmentation. In 2016 fourth international conference on 3D vision (3DV), pages 565–571. IEEE, 2016.

Bram Van Ginneken, Samuel G Armato III, Bartjan de Hoop, Saskia van Amelsvoort-van de Vorst, Thomas Duindam, Meindert Niemeijer, Keelin Murphy, Arnold Schilham, Alessandra Retico, Maria Evelina Fantacci, et al. Comparing and combining algorithms for computer-aided detection of pulmonary nodules in computed tomography scans: theanode09 study. Medical Image Analysis, 14(6):707–722, 2010.

Spyridon Bakas, Mauricio Reyes, Andras Jakab, Stefan Bauer, Markus Rempfler, Alessandro Crimi, Russell Takeshi Shinohara, Christoph Berger, Sung Min Ha, Martin Rozycki, et al. Identifying the best machine learning algorithms for brain tumor segmentation, progression assessment, and overall survival prediction in the brats challenge. arXiv preprint 1811.02629, 2018.

Francisco Silva, Tania Pereira, Julieta Frade, José Mendes Claudia Freitas, Venceslau Hespanhol, José Luis Costa, António Cunha, and Hélder P Oliveira. Pre-training autoencoder for lung nodule malignancy assessment using ct images. Applied Sciences, 10(21):7837, 2020.

Florian Dubost, Hieab Adams, Pinar Yilmaz, Gerda Bortsova, Gijs van Tulder, M Arfan Ikram, Wiro Niessen, Meike W Vernooij, and Marleen de Bruijne. Weakly supervised object detection with 2d and 3d regression neural networks. Medical Image Analysis, 65:101767, 2020.

Valerie W Rusch, Hisao Asamura, Hirokazu Watanabe, Dorothy J Giroux, Ramon Rami-Porta, and Peter Goldstraw. The iaslc lung cancer staging project: a proposal for a new international lymph node map in the forthcoming seventh edition of the tnm classification for lung cancer. Journal of thoracic oncology, 4(5):568–577, 2009.

Olaf Ronneberger, Philipp Fischer, and Thomas Brox. U-net: Convolutional networks for biomedical image segmentation. In International Conference on Medical image computing and computer-assisted intervention, pages 234–241. Springer, 2015.

Kaiming He, Xiangyu Zhang, Shaoqing Ren, and Jian Sun. Deep residual learning for image recognition. In Proceedings of the IEEE conference on computer vision and pattern recognition, pages 770–778, 2016.

Sergey Ioffe and Christian Szegedy. Batch normalization: Accelerating deep network training by reducing internal covariate shift. arXiv preprint 1502.03167, 2015.

Vinod Nair and Geoffrey E Hinton. Rectified linear units improve restricted boltzmann machines. In Proceedings of the 27th international conference on machine learning (ICML-10), pages 807–814, 2010.

Holger R Roth, Le Lu, Ari Seff, Kevin M Cherry, Joanne Hoffman, Shijun Wang, Jiamin Liu, Evrim Turkbey, and Ronald M Summers. A new 2.5 d representation for lymph node detection using random sets of deep convolutional neural network observations. In International conference on medical image computing and computer-assisted intervention, pages 520–527. Springer, 2014.

Mikhail Goncharov, Maxim Pisov, Alexey Shevtsov, Boris Shirokikh, Anvar Kurmukov, Ivan Blokhin, Valeria Chernina, Alexander Solovev, Victor Gombolevskiy, Sergey Morozov, et al. Ct-based covid-19 triage: Deep multitask learning improves joint identification and severity quantification. Medical image analysis, 71:102054, 2021.

